# Autonomic dysfunction characterized by Heart Rate Variability among patients with Sepsis-related Acute Respiratory Failure

**DOI:** 10.1101/2021.11.02.21265811

**Authors:** Preethi Krishnan, Curtis Marshall, Philip Yang, Sivasubramanium V. Bhavani, Andre Holder, Annette Esper, Rishikesan Kamaleswaran

**Author notes:** **Corresponding Author**: Rishikesan Kamaleswaran, Director of Translational Clinical Informatics, Assistant Professor, Departments of Biomedical Informatics, Pediatrics, and Emergency Medicine, Emory University School of Medicine, WMB, 101 Woodruff Circle, Suite 4127, Atlanta, GA. 30322, E.

## Abstract

**Rationale:** To explore the association and implications of using Heart rate variability (HRV) derived from continuous bedside monitoring as a surrogate for detection of Acute Respiratory Failure (ARF) in critically ill sepsis patients.

**Objective:** To analyze HRV measures derived from continuous physiological data captured before ARF-onset to determine whether statistically significant markers can be characterized when compared to sepsis controls.

**Methods:** Retrospective HRV analysis of sepsis patients admitted to Emory Healthcare ICUs was performed between ARF and age and gender-matched controls. HRV measures such as time domain, frequency domain, nonlinear, and complexity measures were analyzed up to 1 hour before the onset of ARF, and a random event time in the sepsis-controls. Statistical significance was computed by the Wilcoxon Rank Sum test.

**Results:** A total of 89 intensive care unit (ICU) patients with sepsis were included in this retrospective cohort study. Time-domain HRV measures including pNN50 (the fraction of consecutive NN intervals that differ by more than 50 ms), RMSSD (root-mean-square differences of successive NN intervals), standard deviation, interquartile range, variance, and approximate entropy for Beat-to-Beat intervals strongly distinguished ARF patients from the controls group. HRV measures for nonlinear and frequency domains were significantly altered (p<0.05) among sepsis patients with ARF compared to controls. Frequency measures such as low frequency (LF), very low frequency (VLF), high frequency (HF), and SD1/SD2 ratio nonlinear measure (SD1:SD2) also showed a significant (p<0.05) increase in the ARF group patients. Multiscale entropy complexity was lower for ARF patients compared to the control counterparts. Detrended fluctuation analysis (DFA) showed a decreasing trend in ARF patients.

**Conclusions:** HRV was significantly impaired across sepsis patients who developed ARF when compared to sepsis controls, indicating a potential prognostic utility for earlier identification of the need for mechanical ventilation and management of patients suspected with sepsis.

## Introduction

Sepsis is defined as life-threatening organ dysfunction caused by a dysregulated host response to infection that is associated with a high mortality rate and significant short-term and long-term morbidity (2–4). One of the major organ dysfunctions that can complicate sepsis is an acute respiratory failure (ARF). The causes of ARF in sepsis include the demand for higher minute ventilation to compensate for the metabolic acidosis, abnormal compliance, and gas exchange due to increased extravascular lung water and pulmonary edema from the capillary leak, and inflammatory lung injury (2). ARF in critically ill patients with sepsis has been associated with prolonged intensive care unit (ICU) stay, which in turn results in increased morbidity and mortality as well as greater expenditure of healthcare resources (5). As such, there is a clinical need to identify and triage patients with sepsis who are at increased risk of developing ARF.

Although the Sequential Organ Failure Assessment (SOFA) and the quick SOFA (qSOFA) scores used in the current Sepsis-3 definition include the partial pressure arterial oxygen to fraction of inspired oxygen (P/F) ratio and respiratory rate (1), these rely on clinical laboratory testing and bedside observation and are not sufficient for identifying ARF associated with sepsis. Heart rate variability (HRV) has been studied as a method to risk-stratify and prognosticate patients with sepsis. HRV measures the oscillations of the intervals between consecutive heartbeats and is a noninvasive and indirect evaluation of the autonomic function. Unlike the P/F ratio and the respiratory rate, HRV is a continuous stream of information that is agnostic to the clinician’s observation or suspicion of a problem. Prior studies have found that HRV can be used to predict not only the risk of developing sepsis, but also several outcomes including the development of septic shock, multiorgan dysfunction, and mortality in patients with sepsis (6). However, HRV has not been studied specifically to predict the risk of developing sepsis-related ARF.

Therefore, in this study, we explore the association and implications of HRV and autonomic dysfunction in critically ill sepsis patients. To test our hypothesis HRV measures were retrospectively compared for the group of patients that developed sepsis-related ARF requiring the need of invasive mechanical ventilation and a sepsis control group that did not.

## Methodology

This retrospective observational cohort pilot study of sepsis-related ARF patients was approved by Emory University Institutional Review Board, IRB #STUDY00000302. Patients admitted between December 2016 to March 2018 within the Emory Healthcare system from the medical, surgical, neurocritical, transplant, and cardiac ICUs were selected to be part of the study. Physiological data from 152 patient beds were collected from Emory affiliated hospitals using the General Electric (GE) bedside monitors and BedMaster system (Excel Medical Electronics, Jupiter FL, USA). The extracted patient data from bedside monitors consisted of multiple days of the continuous high-frequency ECG waveform. The patient data included in the study was exported from the bedside monitoring archive system and de-identified. The patient cohorts were divided into all-cause sepsis patients who met sepsis-3 criteria (1) and developed ARF (requiring invasive mechanical ventilation) and controls were identified as those patients meeting sepsis-3 criteria but did not develop ARF as shown in Fig. 1. The ARF cohort consists of patients with average ARF duration ranging from 6 to 41 hours across different ICU units with a median of 12 hours. Patients who did not meet the sepsis-3 criteria were excluded from this study. The demographic characteristics of ARF sepsis patients and all-cause sepsis patients are described in Table 1.

**Table 1:** Patient demographic characteristics for sepsis-related ARF (Acute Respiratory Failure and sepsis without ARF cohort.

|  | Sepsis-related ARF | Sepsis without ARF<br>(Controls) |
| --- | --- | --- |
| **Patients, n** | 31 | 58 |
| **Valid Data** | 23 (74%) | 58* (100%)* |
| **Age, mean [IQR]** | 55.4 [49.0, 66.0] | 59.1[51.0 70.8] |
| **Age Groups, n**<br>**(column %)** |  |  |
| 18 yrs. to 40 yrs. | 7 (23%) | 8 (14%) |
| 41 yrs. - 60 yrs. | 11 (35%) | 21 (36%) |
| 61 yrs. - 80 yrs. | 12 (39%) | 25 (43%) |
| 81+ yrs. | 1 (3%) | 4 (7%) |
| **Male, number (column %)** | 18 (58%) | 27 (47%) |
| **P/F Ratio, mean (IQR)** | 331.0 [180.8 379.2] | 433.3 [283.9 489.29] |
| **Race, n (column %)** |  |  |
| African American | 17 (55%) | 34 (59%) |
| Caucasian | 11 (35%) | 22 (38%) |
| Asian | 1 (3%) | 1 (2%) |
| Unknown Race | 2 (6%) | 1 (2%) |
| **In hospital deaths, n (column %)** | 12 (39%) | 5 (9%) |

\*For the 24 hours of admit data, 53 (91%) patients had valid data.

**Figure 1:**
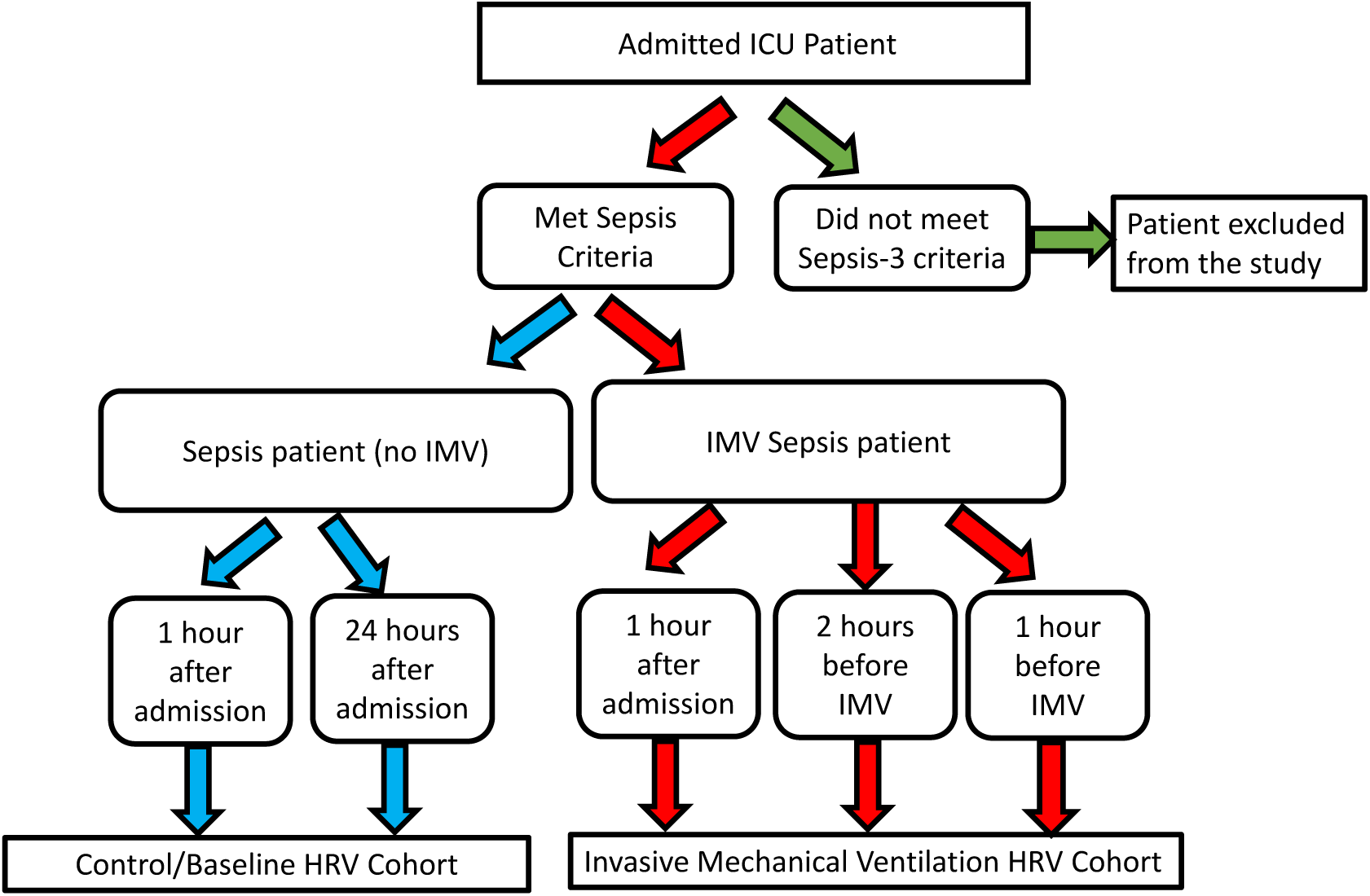
The study population includes all-cause sepsis (controls/baseline) and sepsis-related acute respiratory failure (ARF) patients who underwent invasive mechanical ventilation.

### Data abstraction

Continuous ECGs were captured from the ICU bedside monitor for the ARF and the baseline patient cohorts. ECG waveform was sampled at a sampling frequency of 240 Hertz (Hz) and 23 HRV measures were computed using MATLAB® Physionet Cardiovascular Signal Toolbox (7,8). The HRV measures are described in Table 2. We identified the onset of mechanical ventilation by computing the first presence of a set of ventilator indicator variables (tidal volume, plateau pressure, and PEEP) within the electronic medical record (EMR).

**Table 2:** HRV Features and their descriptions.

| Domain | Parameters | Description |
| --- | --- | --- |
| Time-domain | NNmean | Mean value of NN (peak-to-peak) intervals (ms) |
|  | NNiqr | Interquartile Range of NN intervals |
|  | NNmedian | Median of the NN interval (ms) |
|  | NNmode | Mode of the NN intervals (ms) |
|  | NNvariance | The variance of the NN interval |
|  | NNskew | The skewness of the NN interval |
|  | NNkurt | Kurtosis of the NN interval |
|  | SDNN | The standard deviation of all NN intervals (ms) |
|  | RMSSD | The square root of the mean of the sum of the squares of the NN intervals (ms) |
|  | pnn50 | Count of NN intervals > 50 milliseconds divided by the total number of all NN intervals (%) |
|  | SampEn | Sample Entropy (a.u.) |
|  | ApEn | Approximate Entropy (a.u.) |
|  | PRSA- AC | Acceleration capacity (ms) |
|  | PRSA- DC | Deceleration capacity (ms) |
| Frequency Domain | ULF | Power in the ultra-low frequency range (default < 0.003 Hz) (ms<sup>2</sup>) |
|  | LF | Power in low frequency range (ms<sup>2</sup>) (default 0.04 Hz <= lf < 0.15 Hz) (ms<sup>2</sup>) |
|  | HF | Power in high frequency range (default 0.15 Hz <= hf < 0.4 Hz) (ms<sup>2</sup>) |
|  | LFHF | Ratio LF [ms<sup>2</sup>] / HF [ms<sup>2</sup>] |
|  | TTLPWR | Total spectral power (approximately <0.4 Hz) (ms<sup>2</sup>) |
|  | VLF | Power in very low frequency range (default 0.003 Hz <= vlf < 0.04 Hz) (ms<sup>2</sup>) |
| Non-linear | SD1 | SD1 measure of the Poincaré plot |
|  | SD2 | SD2 measure of the Poincaré plot |
|  | SD1SD2 | SD1/SD2 ratio |

### Case and Control Definition

HRV measures calculated over 300s sliding windows were aggregated over two one-hour periods among the sepsis-related ARF cohort, one immediately at ICU admission, and a second one hour before the onset of invasive mechanical ventilation, defined as ARF. To identify an event time among the control cohort, we selected one hour following ICU admission and a second random one-hour period up to 24 hours post ICU admission. This timeframe was selected to adjust for any variation that may arise due to worsening clinical status as opposed to markers that uniquely predict ARF.

### Personalized Baseline

To evaluate dynamic temporal profiles of HRV among critically ill sepsis patients that progress to ARF, we utilize a personalized baseline approach. In this method, we compute a rolling baseline value for each of the 23 HRV metrics as the Euclidean distance between the current value and the average measure at ICU admission.

### Heatmap Clustering

Each patient data was adjusted using a personalized baseline as described above. The personalized baseline aggregate was log_2_ transformed to characterize increase or decrease in the expression of each HRV metric-i.e., each increase of one represents a doubling in the magnitude of that measure. A heatmap was then generated by applying color-based encoding to the population aggregate of the log-transformed data. Color encoding was scaled to illustrate both increases and decreases in the expression level of the HRV measure relative to the aggregate patient baseline.

### Statistical Analysis

Relevant HRV features were computed using MATLAB® Physionet Cardiovascular Signal Toolbox (7). The statistical analysis of this data was analyzed using Python Scikit-learn (Ref) software. Statistical significance was computed by the Wilcoxon Rank Sum test and one-way ANOVA.

## Results

A total of 89 hospitalized ICU patients with sepsis were included in this retrospective cohort study. Patients who met the sepsis-3 criteria during the ICU stay and a high-frequency ECG waveform were captured from the bedside monitor. From this cohort, 31 patients underwent invasive mechanical ventilation. ECG data one hour after admission, one hour before invasive mechanical ventilation, and two hours before invasive mechanical ventilation were considered for the sepsis-related ARF cohort. The sepsis control cohort consisted of 57 patients who did not develop ARF and consequently did not require mechanical ventilation. Control (8 patients) and ARF (5 patients) with insufficient ECG data or poor quality have been excluded from this study.

The mean age of the mechanical ventilation and the control group did not differ significantly. There were more males in the ARF cohort, and the proportion of African American patients was higher in both groups. There was a higher incidence of sepsis among older patients in both groups. Furthermore, sepsis leading to ARF was correlated with higher mortality as shown in Table 1.

The Uniform Manifold Approximation Projection (UMAP) plot in Figure 2 shows distinct clustering for controls (Figure 2A) and the sepsis-related ARF cohort (Figure 2B). While two visible clusters are observed in both cohorts, greater separation is seen in the feature space between the sepsis-related ARF cohort, suggesting greater dynamics among the HRV features. Notably, the clusters within the ARF cohort reveal a grouping of periods between 0-2 mins before ARF onset (blue highlight region, Figure 2B), and a second cluster containing data about an earlier period containing 5-10 min temporal periods. However, the same temporal regions are organized at random in the control cohort, indicating a possible correlation with physiological deterioration among the ARF cohort.

**Figure 2:**
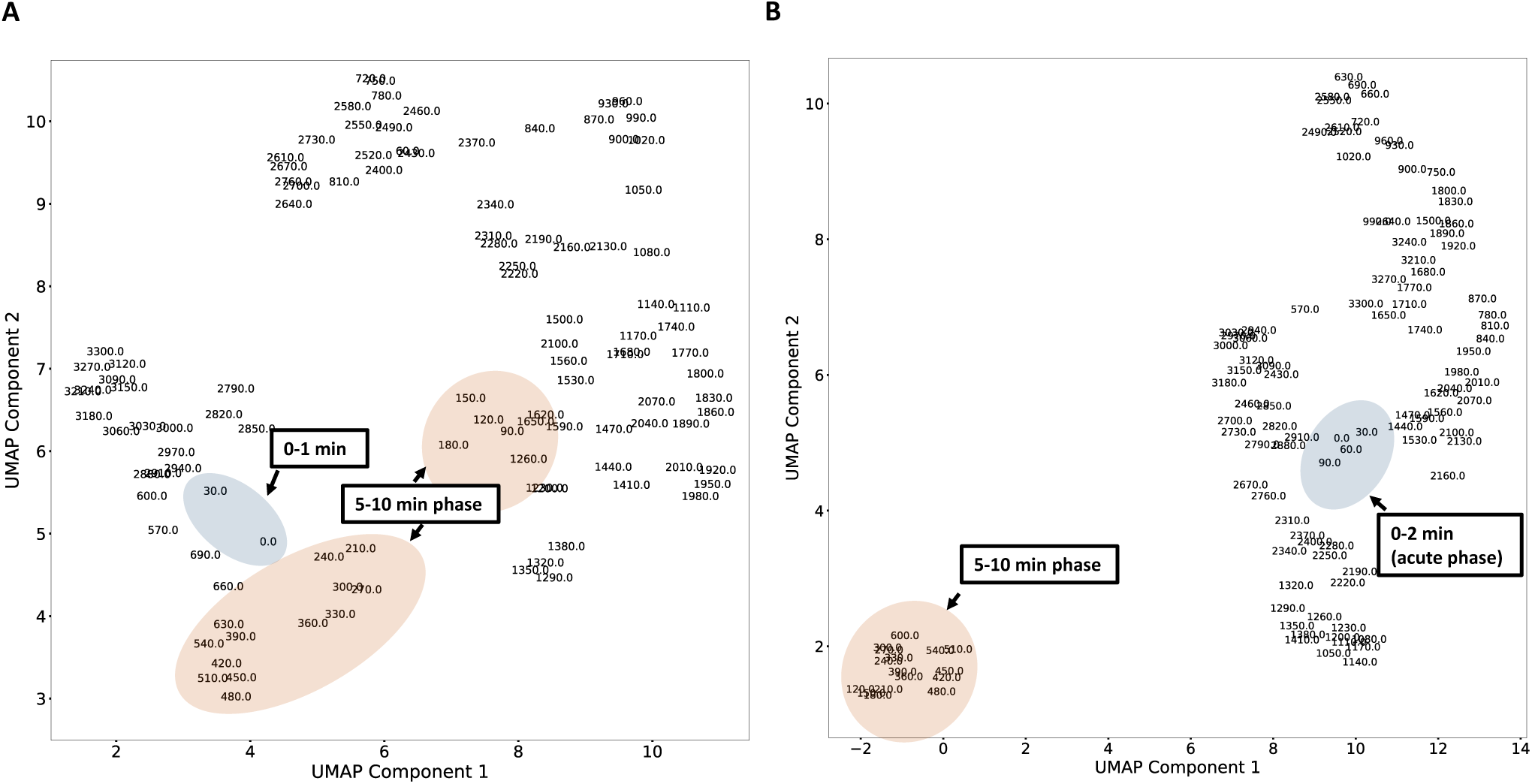
The Uniform Manifold Approximation Projection (UMAP) plot shows distinct clustering for controls and the sepsis-related ARF cohort. A. The controls cohort UMAP plot shows that the temporal regions are organized at random. B. Sepsis-related ARF cohort UMAP suggests greater dynamics among the HRV features. The clusters within the ARF cohort reveal a grouping of periods between 0-2 mins before ARF onset (blue highlight region), and a second cluster containing data about an earlier period containing 5-10 min temporal periods.

Figure 3 illustrates a selected set of time-domain HRV box plots that were statistically significant among sepsis patients with and without ARF. Fig 3A shows a statistically significant increase (P<0.05) in Beat-to-Beat (NN) interval variance among ARF patients one hour before the onset of invasive mechanical ventilation when compared to their baseline at admission. A similar trend is observed in Beat-to-Beat interquartile range (Fig. 3B), standard deviation (Fig. 3B), root mean square of the successive difference between beats (RMSSD - Fig. 3D), and the percentage of absolute differences in successive NN values > 50 ms (pnn50 - Fig. 3E). The Beat-to-Beat interval metrics such as interval variance and standard deviation show significant QRS complex elongation between ARF and control patients. RMSSD and pNN50 measures relate to the vagal tone.

**Figure 3:**
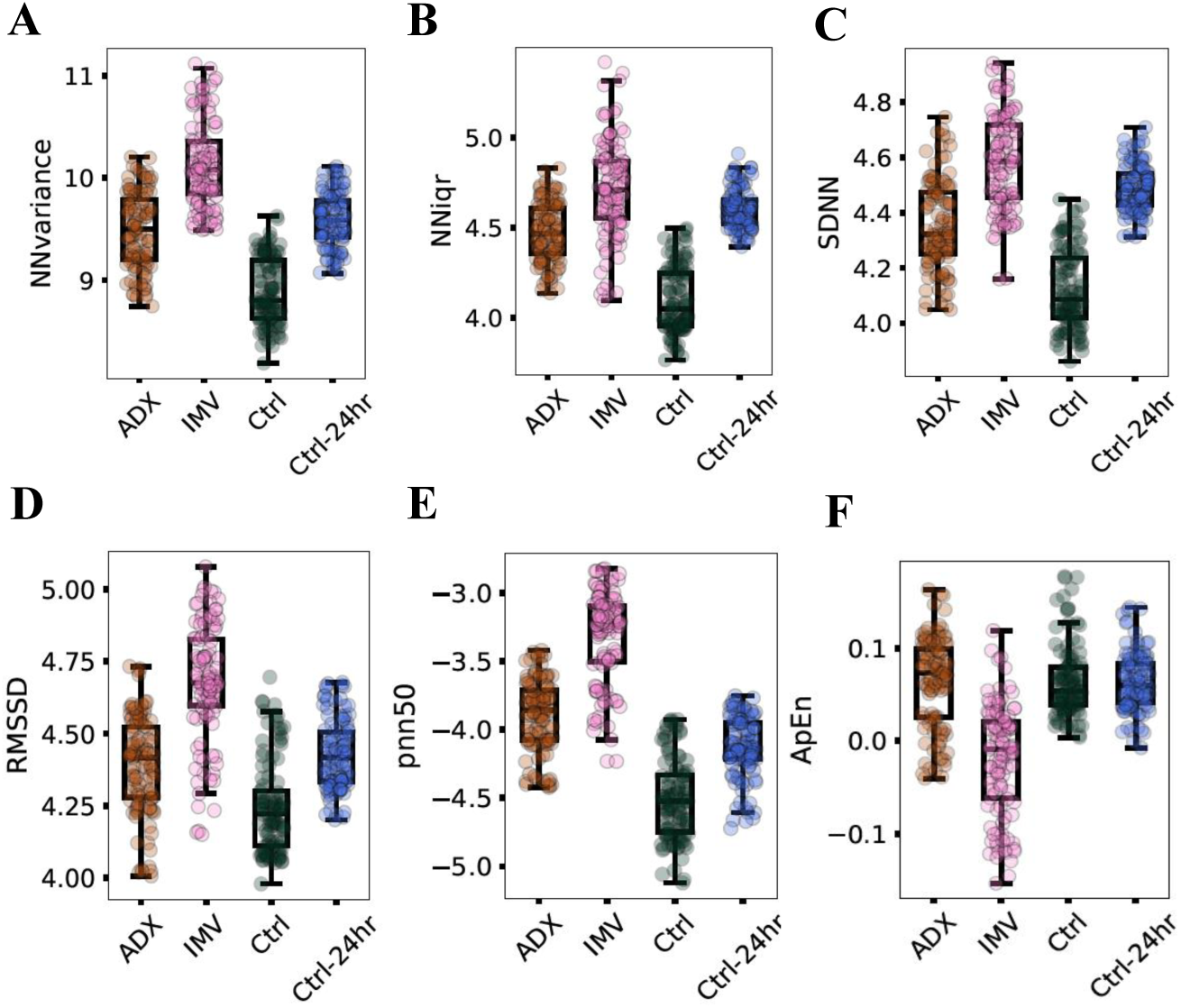
The box-plot panel shows a selected set of time-domain HRV measures that were statistically significant among sepsis acute respiratory failure patients in comparison with all-cause sepsis patients. The controls cohort includes HRV data 1 hour at intensive care unit (ICU) following admission (Ctrl) and 24 hours after ICU admission (Ctrl −24hr). The ARF cohort includes HRV data for 1 hour following ICU admission (ADX) and 1 hour before invasive mechanical ventilation (IMV). The time-domain HRV measures shown in the figure are as follows: A. Beat-to-Beat (NN) interval variance (NNvariance). B. NN interquartile range (NNIqr). C. NN Standard Deviation (SDNN). D. Square root of the mean of the sum of the squares of the NN intervals (RMSSD). E. Count of NN intervals > 50 milliseconds divided by the total number of all NN intervals (pnn50). F. Approximate Entropy (ApEn).

A set of four HRV measures for nonlinear and frequency domain were statistically significant (P<0.05) among sepsis patients with ARF. Frequency measures such as low frequency (LF), very low frequency (VLF), high frequency (HF), and SD1/SD2 ratio nonlinear measure (SD1:SD2) show a statistically significant (p<0.05) increase for ARF patients (Fig. 4A - Fig. 4D). The ratio of LF/HF frequency measure and Approximate Entropy (ApEn) show a statistically significant decrease when compared to the control cohort as shown in Fig. 3F and 4D respectively.

**Figure 4:**
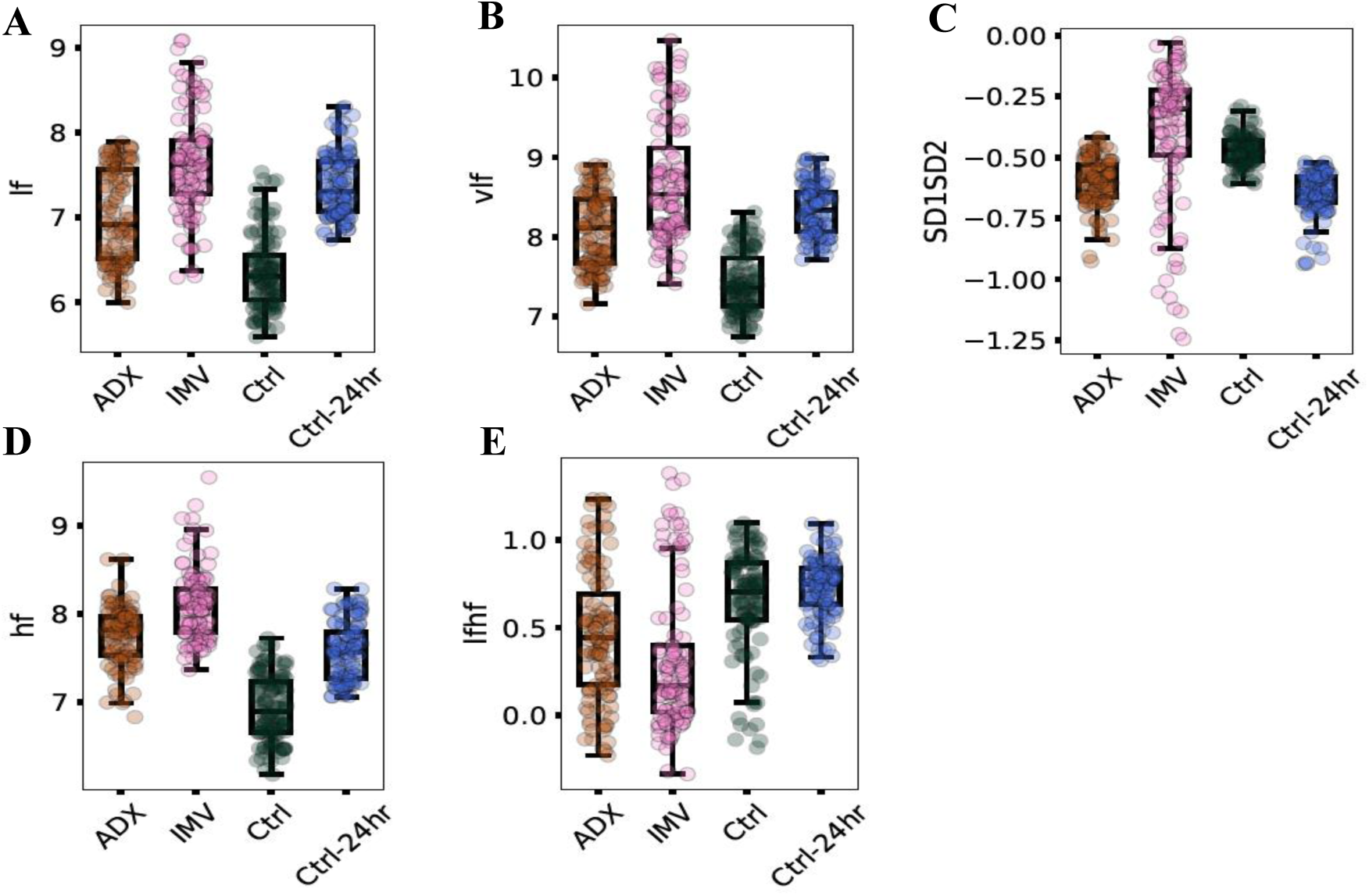
The box-plot panel illustrates a selected set of frequency domain and nonlinear HRV measures that were statistically significant among sepsis acute respiratory failure patients in comparison with all-cause sepsis patients. The controls cohort includes HRV data 1 hour at intensive care unit (ICU) following admission (Ctrl) and 24 hours after ICU admission (Ctrl-24hr). The ARF cohort includes HRV data for 1 hour following ICU admission (ADX) and 1 hour before invasive mechanical ventilation (IMV). The nonlinear frequency domain and nonlinear HRV measures shown in the figure are as follows: A. low frequency (lf). B. very low frequency (vlf). C. SD1/SD2 ratio nonlinear measure (SD1SD2). D. high frequency (hf). E. Ratio LF / HF (lfhf).

Fig. 5 and Fig. 6 illustrate a heatmap as a function of HRV dynamics over time among ARF and controls respectively. Fig. 5 depicts the temporal expression profile of standard HRV metrics in the one hour before the onset of invasive mechanical ventilation. The heatmap characterizes increased and decreased expression using a personalized benchmark that compares the current magnitude of each HRV measure against the associated metric calculated at 1-hour post ICU admission. Several measures, including NN, mean, median and mode significantly increased expression (4 doubling factors) in the period closest to the onset of ARF (e.g., 0-540 seconds). pNN50, a marker of autonomic dysfunction, is seen to be significantly increased in a similar timeframe. Fig. 6 illustrates a similar temporal response among sepsis patients without ARF at 24-hour post ICU admission. NN mean median, mode, and pNN50 were found to be under-expressed relative to the baseline at ICU admission.

**Figure 5:**
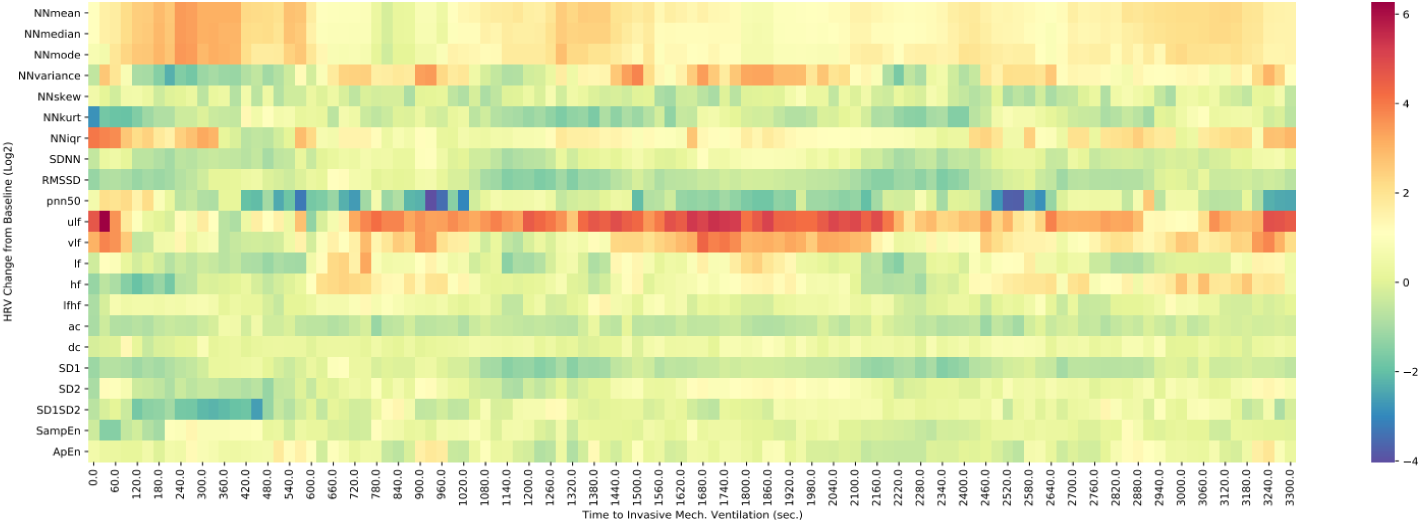
The figure depicts a heatmap of HRV statistical measures over time among ARF temporal response in the one hour before the onset of invasive mechanical ventilation (IMV). NN, mean, median, mode, and pNN50 are seen to be significantly increased one hour before mechanical ventilation.

**Figure 6:**
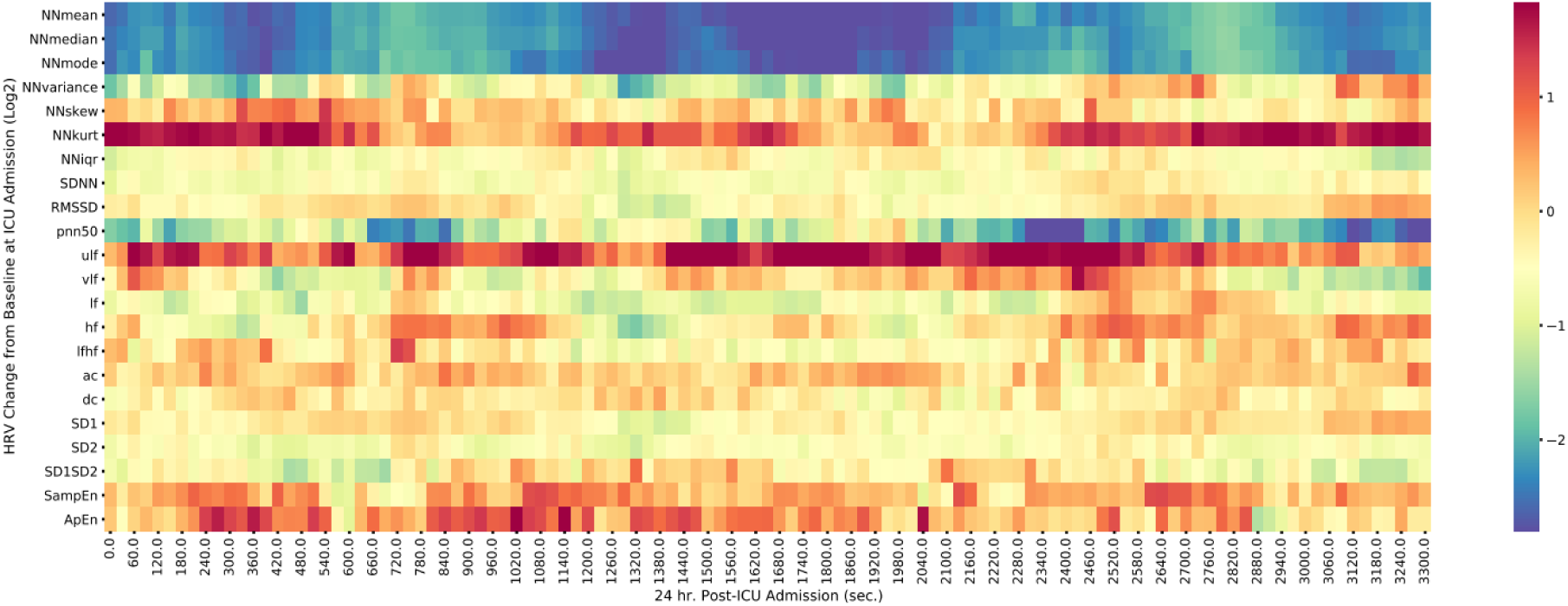
The figure depicts a heatmap of HRV statistical measures over time among sepsis patients without ARF at 24-hour post ICU admission. NN mean median, mode, and pNN50 were found to be under-expressed relative to the baseline at ICU admission.

MSE (Multiscale Entropy) and DFA heart rate complexity analyses were performed for the control and sepsis-related ARF groups. Fig. 7 shows a multiscale entropy (MSE) heart rate complexity analysis plot for the controls and sepsis-related ARF group. The MSE plot includes mean values for sepsis-related ARF patients one hour after ICU admission and 2 hours before invasive mechanical ventilation. The controls sepsis patient’s mean values include data one hour after ICU admission and 24 hours after ICU admission. While select time scales (6, 7, 9, 10) in the control groups significantly increased between admission and 24 hours (p<0.05), this was abolished in the ARF cohort. Patients who would eventually need a ventilator started at a lower baseline entropy compared to the control counterparts. On the other hand, the increase in entropy in the ARF group shortly before initiation of ventilation was not as high as the controls. The upper and lower 95% confidence interval of the mean entropy values for time scales (6,7, 9, 10) are in the range (0.94-1.36) and (0.62-0.96). The DFA analysis was not statistically significant (p>0.05) for sepsis patients of both groups. Table 3 shows a decrease in mean DFA α1 and DFA α2 for ARF patients in comparison to control values 24 hours after ICU admission.

**Table 3.** Detrended fluctuation analysis (DFA) mean values.

| **Patient Group** | **DFA $\alpha 1$**<br>**(Mean)** | **DFA $\alpha 2$**<br>**(Mean)** |
| --- | --- | --- |
| Controls - 1 hour after admission | 0.71 | 0.96 |
| Controls – 24 hours after admission | 0.79 | 0.97 |
| ARF – 1 hour after admission | 0.75 | 1.02 |
| ARF – 2 hours before mechanical ventilation | 0.77 | 0.94 |

ARF – Acute Respiratory Failure

**Figure 7:**
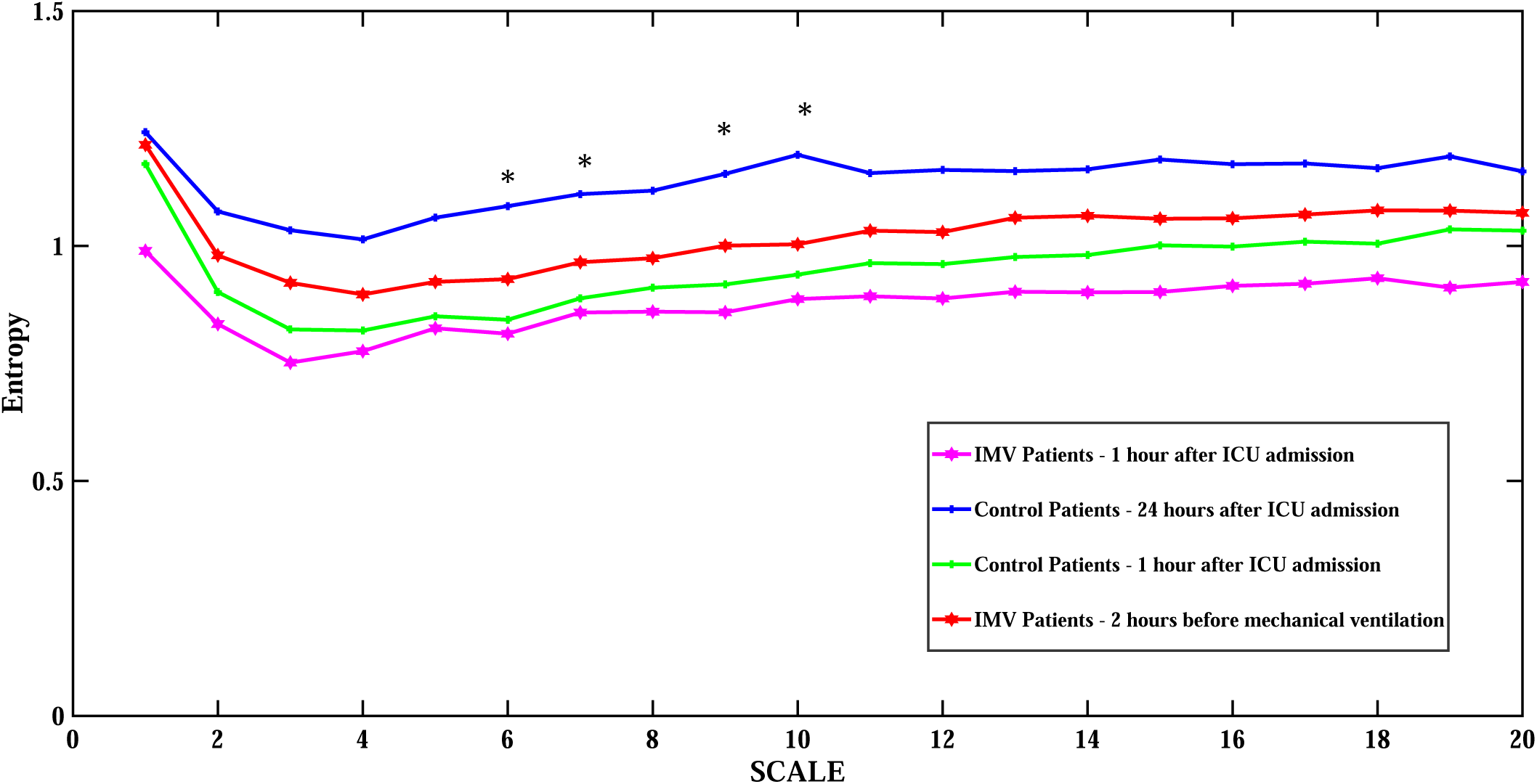
The figure shows a multiscale entropy (MSE) heart rate complexity analysis plot for the controls and sepsis-related ARF group. The MSE plot includes mean values for sepsis-related ARF patients one hour after ICU admission and 2 hours before invasive mechanical ventilation. The controls sepsis patient’s mean values include data one hour following ICU admission and 24 hours after ICU admission. The timescales for the control cohort when p<0.05 are denoted by the * symbol.

## Discussion

This study has two major findings. First, our results show that HRV measures including time domain, frequency domain, nonlinear and complexity measures are associated with autonomic dysfunction for sepsis-related ARF patients. Second, HRV as a non-invasive measurement of the autonomic nervous system (ANS) function may be used to evaluate critically ill sepsis-mediated ARF patients’ need for invasive mechanical ventilation.

The leading causes of ICU admission are sepsis and ARF (9). The treatment for acute respiratory failure involves providing mechanical ventilation and is dependent on the balance between oxygen supply and uptake (10). HRV analysis is a non-invasive technique and is a readily obtainable measure at the patient bedside. It has been used to detect clinical deterioration and increased mortality among critically ill sepsis patients (6). By comparing retrospective cohorts of sepsis ARF patients and sepsis controls, this pilot study demonstrates distinctively expressed subsets of markers that predict the onset of ARF by utilizing time domain, frequency domain, nonlinear, and complexity HRV measures. The results further show statistical separation (p<0.05) across several HRV measures (Fig 3, Fig 4, Fig 6) between sepsis patients with ARF and all-cause sepsis patients.

Retrospective HRV data used for the study was one hour and 24 hours post-admission for the all-cause sepsis control patients. The interval of 24 hours was considered to avoid recording any possible artifacts that might have been generated during ICU admission. ARF patient retrospective HRV data one hour after admission, 1 hour, and 2 hours before mechanical ventilation was analyzed for this study.

Among the significant time-domain HRV measures (Table 2), pNN50, RMSSD, standard deviation, interquartile range, variance, and approximate entropy for NN intervals strongly distinguished ARF patients from the controls group. pNN50 represents the proportion of pairs of successive NN intervals that differ by more than 50 ms and have been associated with poor autonomic health and an increased risk of mortality. In addition, pNN50 is influenced by respiration, to be a predictor of congestive heart failure, and is also a marker of parasympathetic function (11). In this study, pNN50 (Fig 3E) provides useful information about sinus rhythm dynamics in health and disease, related to parasympathetic regulation.

SDNN is a commonly used temporal HRV parameter that may be associated with sepsis mortality and reflect all the cyclic components responsible for HRV (6). SDNN regulates the autonomic function in a parasympathetically mediated manner, and the values are influenced by age and associated ANS function. A greater value of NN variance (Fig. 3A and Fig. 3B) corresponding to a lower heart rate and interquartile range is a predictor of mortality. SDNN is also a prognostic indicator of cardiovascular risk in different populations (12). An increased RMSSD (Fig. 3D) is prevalent in septic shock and could contribute to a potential increase in risk for mortality. (13). For normal sinus rhythm and normal AV-nodal function, RMSSD and pNN50 contribute to parasympathetic modulation of normal R-R intervals driven by ventilation (11). Approximate entropy is a measure for complex data and identifies changes in time series by assigning a non-negative number to the series. IMV patients showed a lower approximate entropy (Fig. 3F) than the controls group, which indicates a decrease in the heart rate modulation dynamics. Poor heart rate dynamics result in impairment of lung function, and impaired lung diffusion capacity is related to an altered parasympathetic response (14).

The ANS regulates blood pressure, heart rate, and respiration by cardiovascular control between the heart, lungs, and brain. ICU patients have a high incidence of ANS dysfunction leading to an increase in poor clinical outcomes and mortality (15). Predictive scoring systems such as APACHE II (Acute Physiology and Chronic Health disease Classification System II) and SOFA (Sepsis-related Organ Failure Assessment) do not consider composition changes in the ANS (6). The parasympathetic HRV variables, RMSSD, pNN50 are correlated with the HF band and influence vagal activity.

LF/HF HRV measures indicate both ventilatory modulations for R-R intervals (RSA - Respiratory Sinus Arrhythmia) with the efferent impulses on the cardiac vagus nerves and baroreflexes with a combination of sympathetic, parasympathetic efferent nerve traffic to the sinoatrial node (11). Thus, the frequency bandwidths LF and HF can be correlated with autonomic nervous system (ANS) regulation of sympathetic, and parasympathetic activity. During acute inflammation, there is an increase in LF (Fig. 4A) to maintain physiological homeostasis. There is also an increase in HF (Fig. 4D) due to the ANS response to infection-related endotoxin-mediated inflammation. The ratio of the two bands (LF/HF) represents sympathovagal balance (16). A decreased LF/HF ratio (Fig. 4E) due to high HF may be considered as a detection index for either sympathovagal balance or sympathetic modulations. LF, HF, LF/HF may be used as a metric to predict malignant ventricular arrhythmias, cardiac death, and arrhythmic death. VLF power reflects the activity of the renin-aldosterone system, thermoregulation, or vasomotor activity. ULF (Ultra-low frequency) and VLF (Fig. 4B) are powerful risk predictors in cardiovascular diseases. ULF power had the strongest association with these fatal outcomes (11). ULF, a marker of slow-onset biological processes, such as changes in core body temperature and circadian rhythm is shown to be significantly elevated in both ARF and the control cohorts for much of the observational window, suggesting possible pathological dynamics associated with sepsis rather than being specific to ARF.

Cardiopulmonary Coupling (CPC) can be calculated using HRV and respiratory information. HRV and CPC methods may be used to assess mechanical ventilation weaning readiness. RSA improves the pulmonary gas exchange and cardiac efficiency through ANS, vagus nerve together with mechanical effects, and the baroreflex. Both respiration and HRV have a causal relationship as respiration drives acceleration or deceleration in the heart rate through RSA (15). Nonlinear HRV measures quantify the structure of the NN time series which cannot be ascertained using the time and frequency domain measures and show the relationship between hemodynamic, electrophysiological, humoral variables, autonomic, and central nervous system (13). SD1 is a nonlinear measure which is the standard deviation measuring the dispersion of points in the plot perpendicular to the line of identity. SD2 is the standard deviation measuring the dispersion of points along the line of identity (14). SD1SD2 is derived from the Poincaré Plot and is a measure of autonomic balance. Poincaré plots identify abnormal NN intervals, and an extremely low HR is referred to as abnormal. An increased SD1SD2 as shown in Fig. 4C may indicate broad and complex dynamics between the sympathetic and parasympathetic arms. Nonlinear metric SD1 measures correlate with baroreflex sensitivity and predict diastolic BP. While RMSSD is identical to the non-linear metric SD1, SD2 correlates with LF power, and SD1/SD2 is correlated with the LF/HF ratio. The ratio of SD1/SD2 measures unpredictability and is used to measure autonomic balance when there is sympathetic activation (17).

MSE and DFA measure the complexity of the underlying heart rate dynamics due to complex connections among the autonomic, endocrine, and cardiovascular systems. A lower MSE and DFA for ARF patients as shown in Fig. 7 and Table 3 indicates that as a patient disease progresses, there is an associated decrease in complexity values (18). Individual MSE scale values may help in ARF detection and prognosis. Lower DFAα1 below 0.95 has been shown to significantly increase non-cardiac mortality related to sepsis. Sympathetic over-activation may play an important role in the increased mortality in the ARF group and could be detected by DFA (19).

HRV time-domain markers such as NN variance, NN IQR, SDNN, and RMSSD in IMV patients showed an increase when compared to control patients, which are opposite to what is observed in the sepsis literature. A similar increase is seen in frequency domain HRV measures. The increase suggests complex dynamic mechanisms of autonomic dysfunction, electrophysiology, and hemodynamic systems in sepsis ARF patients. In conclusion, these results indicate that HRV measures such as time, frequency, nonlinear domain, and MSE complexity analysis may be used as noninvasive physiological biomarkers to predict the need for invasive mechanical ventilation in critically ill sepsis patients associated with ARF.

### Limitations

A smaller group of sepsis patients (89) from a single urban academic center was considered for this study. ICU patients with no usable ECG recordings were excluded. Factors such as body position, respiratory rate, and mechanical ventilation weaning can influence HRV measures. ARF recovery outcomes influenced by the above factors were not part of the HRV analyses. Pharmacological interventions such as vasopressors and the influence of comorbidities for ARF will be evaluated in our future work.

## Data Availability

All data produced in the present study are available upon reasonable request to the authors

## Acknowledgment

RK was funded by NIH NIGMS under R01GM139967, and NCATS under UL1TR002378 and the Surgical Critical Care Initiative (SC2i), Department of Defense Health Program Joint Program Committee 6 / Combat Casualty Care (USUHS HT9404-13-1-0032 and HU0001-15-2-0001). PY is supported by National Heart, Lung, and Blood Institute (NHLBI 5T32HL116271-08). SVB is supported by the National Institute for General Medical Sciences of the National Institutes of Health (K23GM144867). ALH is supported by the National Institute of General Medical Sciences of the National Institutes of Health (K23GM37182). The content is solely the responsibility of the authors and does not necessarily represent the official views of the National Institutes of Health.

